# Diagnostic accuracy of physical examination to detect the pelvic fractures among the blunt trauma patients; systematic review and meta-analysis

**DOI:** 10.1101/2020.01.09.20017129

**Authors:** Yohei Okada, Norihiro Nishioka, Yasushi Tsujimoto

## Abstract

**Background:** The study aims to perform systematic review and meta-analysis to identify the diagnostic accuracy of physical examination for pelvic fracture among the blunt trauma patients.

**Method:** We will perform a systematic review and meta-analysis for diagnostic test accuracy (DTA). We will include all reports on the diagnostic accuracy of physical examinations for detecting pelvic fractures. We will include the studies designed as prospective or retrospective observational (cohort or cross-sectional) studies or secondary analysis of randomized controlled trials. The target participants are blunt trauma patients with potential pelvic injury. The target condition is pelvic fracture. The index test being investigated is physical examination for pelvic fracture. The reference standard is X-ray or computed tomography to confirm the target condition. We will search MEDLINE, EMBASE and The Cochrane Library inclusive of Cochrane Controlled Trials Register. Two authors will independently screen the study eligibility and extract data. Screening will be a two-step process with initial title/abstract screening followed by full-text screening. We will evaluate the risk of bias independently by two investigators and reported according to the QUADAS-2 tool. In the meta-analysis, we will use a bivariate random-effects model to report the summary receiver operating characteristic (SROC) point (summary values for sensitivity and specificity) and the 95% confidence region around the summary ROC point.

**Trial registration:** This review is submitted with University hospital medical information network clinical trial registry (UMIN-CTR) [UMIN000038785].

## Background

### Target condition being diagnosed

Pelvic fracture is a life-threatening injury which cause retroperitoneum hemorrhage and hemorrhagic shock among blunt trauma patients.^1-3^ Recent reports indicated ten to fifteen percent of patients with pelvic fractures were in shock on arrival to the ED, and one-third of them died.^4^ For saving such a patient, it is essential to recognize the injury and to start intervention as soon as possible. ^1 2^

### Clinical pathway

Physical examination of pelvis by paramedics or physicians is to detect the pelvic fracture for blunt trauma patients. The examination includes inspection of deformity, simultaneously palpating both iliac crests on either side of the pelvis and applying internal and external rotational stress, anteroposterior and superior-inferior stress. ^3 5^ Especially, it may be useful as a triage in the settings where X-ray or other imaging modalities are unavailable (i.e., at the scene of injury, in the transport modality such as ambulance or helicopter, primary care settings or resource-limited environment). If pelvic fracture is suspected by the examination, temporary fixation of the pelvis by pelvic binder is recommended to prevent the further bleeding, and they should be transferred to tertiary care centers for definitive diagnosis by x-ray and/or Computed tomography (CT) and the treatment.^1^

### Why perform this review

Physical examination of pelvis is expected to detect pelvic fracture earlier and enable us to judge the severity and necessity for more accurate diagnosis modality such as x-ray or CT, and to prepare the earlier intervention. However, the false-negative of physical examination may lead to underestimate the severity of injury, and the delay to start appropriate intervention. On the other hand, false-positive may increase the radiation exposure and costs using unnecessary x-ray and/or CT. Therefore, it is necessary to identify the diagnostic ability of physical examination for the pelvic fracture.

Although previous meta-analysis^6 7^ reported pooled diagnostic abilities, these had important limitations. In the first review published in 2004 ^6^, quality assessment was thought as insufficient due to absence of systemic Quality Assessment of Diagnostic Accuracy Studies (QUADAS-2) tool.^8^ Further, these reviews failed to incorporate several important studies due to the inadequate or out-of-date search ^9-14^. We, therefore, perform a systematic review to include all relevant studies following rigorous methodological guidelines.^8 15 16^

### Objective

The study aims to perform systematic review and meta-analysis to identify the diagnostic accuracy of physical examination for pelvic fracture among the blunt trauma patients.

## Method

We will perform a systematic review and meta-analysis for diagnostic test accuracy (DTA). We will adhere to standards of the methodology of the Cochrane handbook for DTA^15^ and the Preferred Reporting Items for a Systematic Review and Meta-analysis of Diagnostic Test Accuracy Studies (The PRISMA-DTA Statement)^16^ in reporting the findings of this review. The review protocol is available in the pre-print server (medRexiv). This review is submitted with the International Prospective Register of Systematic Reviews (PROSPERO) and University hospital medical information network (UMIN) clinical trial registry [UMIN000038785].^17 18^

### Criteria for considering studies for this review

#### Type of studies

We will include all reports on the diagnostic accuracy of physical examinations for detecting pelvic fractures. We will include the studies designed as prospective or retrospective observational (cohort or cross-sectional) studies or secondary analysis of randomized controlled trials. We will exclude diagnostic case-control studies. We will exclude case studies that did not provide sufficient diagnostic test accuracy data, namely true-positive (TP), false-positive (FP), true-negative (TN), and false-negative (FN) values, based on the reference standard.

#### Participants

The target participants are blunt trauma patients with potential pelvic injury.

#### Target condition

The target condition is pelvic fracture due to blunt trauma.

#### Setting

Any type of settings are included. (e.g., pre-hospital setting or emergency department)

#### Index test

The index test being investigated is physical examination for pelvic fracture. Based on the previous literature and standard guidelines for trauma care ^3 5-7^, the positive physical examination is defined as follows: the presence of any pelvic deformity, hip dislocation, ecchymosis, laceration, hematoma over the pelvic ring in inspection, pelvic bony pain or tenderness in palpating, instability or abnormal movement in applying manual internal and external rotational stress, anteroposterior and superior-inferior stress. We also consider positive physical examination as the authors of primary studies defined; however, we will check the robustness by excluding different definition of index test positive in the sensitivity analysis.

#### Reference standard

The reference standard is X-ray or Computed tomography (CT) to confirm the target condition defined as fracture of pelvis (ilium, ischium, pubis, and sacral fractures) diagnosed by physicians such as emergency physicians, acute care surgeons, orthopedics or radiologists. We also consider positive physical examination as the authors of primary studies defined; however, we will check the robustness by excluding different definition of reference standard in the sensitivity analysis.

### Search methods for identification of studies

#### Electronic searches

An electronic search strategy has been developed in collaboration with Kyoto University Medical Librarian. To identify all prospective, retrospective or RCTs, we will search MEDLINE, EMBASE and The Cochrane Library inclusive of Cochrane Controlled Trials Register. We will search for international clinical trials registry platform (ICTRP) and ClinicalTrials.gov to find ongoing and unpublished studies. There are no limits regarding language and publication date for this review. The search strategy is described in supplementary. Also, we will hand-search reference lists of relevant articles.

### Data collection and analysis

#### Selection of studies

Two authors will independently screen the study eligibility and extract data. Screening will be a two-step process with initial title/abstract screening followed by full-text screening. Disagreements among reviewers will be resolved through consensus or third-party reviewer. Following full-text screening, a list of excluded studies with reasons for exclusion will be provided in an appendix of the final report.

#### Data extraction and management

Two authors will develop the data extraction sheet including the following information.

✓ Study characteristics: author, year of publication, country, design, sample size, clinical settings, number studied, and funding source.
✓ Population characteristics: inclusion/exclusion criteria, number of drop-outs with reason and patient demographics such as age and sex.
✓ Index test: timing of sampling, method of examine, time to result, and who performs it.
✓ Reference standard: timing of sampling, method of examine, time to result, and who performs it.
✓ Outcomes: From the 2 × 2 table, we will calculate true positives, false positives, true negatives, and false negatives.

#### Assessment of methodological quality

We will evaluate the risk of bias independently by two investigators and reported according to the QUADAS-2 tool.^8^ A statistical assessment of publication bias will not be performed. There is no evidence of publication bias in systematic reviews of diagnostic accuracy, and methods for detecting publication bias are unreliable when applied to diagnostic accuracy studies. We, therefore, look at the number of ongoing and unpublished studies for the assessment of publication bias.

#### Statistical analysis and data synthesis

We will describe the included available studies summarized in two tables. The first table will summarize the study designs, participants, index test details, sample types, and the reference standards. The second one will summarize the details on study quality relating to QUADAS-2.

The results data of physical examination will be compared to the reference (X-ray or CT). Data for 2 × 2 tables of physical examination against reference standard will be extracted from each study. The true positive, true negative, false positive and false negative will be calculated. If these data are not provided, they will be calculated from raw data if possible. A summary table of evidence will be produced, and individual studies represented using forest plots displaying the sensitivity and specificity values of the physical examination with 95% confidence intervals in order to inspect between-study variability. We will also describe accuracy estimates of individual studies using a receiver operating characteristic (ROC) plot of sensitivity versus 1-specificity to visually assess the correlation between both indices.

In the meta-analysis, we will use a bivariate random-effects model to report the summary receiver operating characteristic (SROC) point (summary values for sensitivity and specificity) and the 95% confidence region around the summary ROC point. All analyses will be performed using SAS, R software (version 1.1.456; R Studio Inc.) or Review Manager 5.3 (Cochrane Collaboration, London, United Kingdom).

#### Investigations of heterogeneity

We will perform subgroup analyses on the following groups if available.

✓ Age (Elderly/Adult/children)
✓ The condition of the patients (such as coma or shock)
✓ Settings (Pre-hospital, in-hospital, or others)
✓ The type of clinicians (Physicians, paramedics, experience or other type)
✓ The type of pelvic fracture (Unstable*/Stable)
✓ The type of reference standard (X-ray/ CT)

*Unstable pelvic fracture: World Society of Emergency Surgery gradeIV^1^

#### Sensitivity analysis

We will assess the robustness by excluding the different definition of index test positive or standard reference.

## Discussion

This systematic review and meta-analysis will evaluate the diagnostic accuracy of physical examination to detect the pelvic fracture. This review has some potential strengths compared with previous meta-analysis. First, it is expected to include all potential literature to estimate the diagnostic accuracy based on the comprehensive literature search. Secondly, we will systematically assess the quality of the included studies. Thirdly, we will perform the subgroup analysis to identify the heterogeneity of diagnostic accuracy. If the heterogeneity exists, we can find which conditions or situations the physical examination should be applied or not.

Further, we describe the potential clinical implication of this review. If the sensitivity of the physical examination is estimated as high, it may be useful as triage in the pre-hospital or primary care settings. In such a case, it enables us to choose the appropriate candidate for transfer to tertiary centers, and to prepare earlier definitive treatment such as trans-arterial embolization. According to that, the physical examination may potentially contribute to increase the survival chance. If it is low, we will find the unreliability of the physical examination. In such a case, we should pay attention to missing injuries or underestimation of the injury, which may lead to inappropriate selection of hospital and delay to the treatment.

## Data Availability

Not applicable

## Founding

This research did not receive any specific grant from funding agencies in the public, commercial, or not-for-profit sectors.

## Ethics approval and consent to participate

The need for ethical approval and consent was waived for this systematic review.

## Supplementary

### Search strategy (MEDLINE via Ovid)

MEDLINE Ovid (Ovid MEDLINE(R) Epub Ahead of Print, In-Process & Other Non-Indexed Citations, Ovid MEDLINE(R) Daily and Ovid MEDLINE(R) 1946 to Present)

### Population: Trauma patients

1: exp “Wounds and Injuries”/OR trauma.ti,ab. OR injur*.ti,ab.

### Index test: Physical examination

2: exp Physical examination/OR ((physical or clinical) ADJ2 (diagnosis or sign* OR symptom* OR assessment or finding* OR evaluat* or examination*)).ti,ab,kw

### Target condition: Pelvic fracture

3: ((fracture* or disrupt* or displac* or injur* or traum* or rupture*) adj2 (pelvi* or ilia* or pubi*)).ti,ab.

### Exclusion: Review, case report, and animal studies

4: Review.pt OR case reports.pt OR (exp animals/ NOT exp humans/)

5: (1 AND 2 AND 3) NOT 4

### Search strategy (EMBASE)

S1 EMB.EXACT.EXPLODE(Injury)

S2 ab(trauma) OR ti(trauma)

S3 ab(injur*) OR ti(injur*)

S4 S1 OR S2 OR S3

S5 EMB.EXACT.EXPLODE(physical examination)

S6 ab((physical or clinical) N/2 (diagnosis or sign* or symptom* or assessment or finding* or evaluat* or examination*)) OR ti((physical or clinical) N/2 (diagnosis or sign* or symptom* or assessment or finding* or evaluat* or examination*))

S7 ab((fracture* or disrupt* or displac* or injur* or traum* or rupture*) N/2 (pelvi* or ilia* or pubi*)) OR ti((fracture* or disrupt* or displac* or injur* or traum* or rupture*) N/2 (pelvi* or ilia* or pubi*))

S8 S6 OR S5

S9 S8 AND S7 AND S4

